# Progress Update for the Multisite Optical Genome Mapping Evaluation and Validation Study: Prenatal Applications

**DOI:** 10.1101/2023.12.22.23300469

**Authors:** Brynn Levy, Jie Liu, M Anwar Iqbal, Barbara DuPont, Nikhil Sahajpal, Monique Ho, Jingwei Yu, Sam J. Brody, Heather Mason-Suares, Mythily Ganapathi, Aleksandar Rajkovic, Teresa A. Smolarek, Reha M. Toydemir, Peter Bui, Ravindra Kolhe, Roger E. Stevenson

## Abstract

Optical genome mapping (OGM) is an emerging technology with great potential for prenatal diagnosis. OGM can identify and resolve all types of balanced and unbalanced cytogenomic abnormalities in a single test, which are typically assessed by multiple standard of care (SOC) methods including karyotyping, fluorescence in situ hybridization and chromosomal microarray.

To assess OGM’s viability as an alternative to conventional SOC testing, a comprehensive clinical research study was conducted across multiple sites, operators, and instruments to evaluate its accuracy and clinical utility. This report provides an update for the phase 2 results of the ongoing multisite evaluation and validation study evaluating OGM for prenatal applications. In phase 1, 123 prenatal cases were assessed by OGM, and in phase 2, 219 retrospective and prospective prenatal cases have been evaluated. For 71% of cases, at least two SOC tests were performed. The study found that OGM had an overall accuracy of 99.6% and positive predictive value of 100% when compared to all cytogenetic SOC results.

With its standardized workflow, cost-effectiveness, and high-resolution cytogenomic analysis, OGM shows great promise as an alternative technology that uses a single assay to consolidate the multiple SOC tests usually used for prenatal cytogenetic diagnosis.

## Introduction

Prenatal diagnostic testing during pregnancy typically involves analyzing amniotic fluid, chorionic villus samples (CVS), or fetal cord blood. These tests are recommended when there is a positive or unclear result from non-invasive prenatal testing (NIPT), positive maternal serum screen, abnormal ultrasound findings or increased genetic risk due to a family history of a chromosome abnormality. While chromosomal microarray is recommended as first-tier prenatal diagnostic testing to detect pathogenic copy number variants (CNVs), it is still common practice to use multiple methodologies to confirm a final diagnosis. This approach, however, is costly, time-consuming, and labor-intensive.^1–4^

When considering prenatal diagnostic options, it’s essential to weigh the strengths and limitations of each technology. In practice, a combination of tests is often used, either simultaneously or sequentially. For instance, to quickly obtain results after an abnormal NIPT, a fluorescence in situ hybridization (FISH) panel and karyotyping is employed for confirmation. If NIPT is negative, FISH with karyotyping and CMA may be pursued. If a positive FISH or CMA result is obtained, karyotyping/targeted FISH may still be necessary to determine the specific nature of the anomaly, particularly in cases where there may be concerns about potential reproductive risks, such as suspected translocations where a parent could be a carrier.^1^

Optical genome mapping (OGM) represents an emerging cytogenomic technology that provides a cost-effective, comprehensive assessment of various pathogenic cytogenomic variants, including structural variations (SVs) and CNVs at the resolution comparable to CMA.^5–7^ OGM operates by fluorescently labeling ultra-high molecular weight DNA (UHMW DNA) at specific 6-base-pair motif sites (CTTAAG), which are distributed roughly every 6,000 base pairs across the genome. These labeled molecules are then compared to a reference genome to identify genomic abnormalities, ranging from as small as 500 base pairs to as large as entire chromosomal aneuploidies or triploidy.^7–9^

Recent studies have demonstrated a high concordance between OGM and traditional cytogenetic techniques in both constitutional prenatal and postnatal applications.^7–10^ The main objective of this study is to conduct a multisite assessment of OGM’s performance, robustness, and reproducibility in prenatal samples when compared to standard of care (SOC) technologies. Phase 1 of the study, as detailed in Stevenson et al., 2022, drew from 200 data points sourced from 123 distinct prenatal cases.^11^ Building upon this groundwork, this report describes the advancements made in phase 2 of the study, encompassing 219 unique retrospective and prospective cases out of a total of 500 unique cases planned for the entire study. This expanded dataset allows for a more comprehensive evidence-based understanding of the OGM’s progression towards integration as a cytogenomic diagnostic technique.

## Methods

This double-blinded study implemented standardized analysis and interpretation workflow for the classification of SVs detected by OGM across multiple sites, to determine the variability in SV reporting, which is a common phenomenon observed among the participating sites while reporting CNVs from CMA.

### IRB disclosures

IRB-00007527 – University of Rochester Medical Center Office for Human Subject Protection

IRB-AAAT9083 – Columbia University Irving Medical Center, New York, NY, USA

IRB-2022-0865 – Cincinnati Children’s Hospital Institutional Review Board, Cincinnati, OH, USA

IRB-20212956 – Bionano Genomics Inc., San Diego, CA, USA

IRB-22-37290 – University of California San Francisco, San Francisco, CA, USA

IRB-A-#00000150 (HAC IRB # 611298) - Medical College of Georgia, Augusta University, Augusta, GA, USA

### Cohort design & Case Selection

Samples were given anonymous study identification (ID) numbers (i.e.: BNGOSS-xxxx). Retrospective and prospective remnant clinical samples were obtained from individuals who underwent NIPT. Cultured amniocyte or chorionic villus sampling cells were then used for extraction of UHMW DNA for the subsequent OGM workflow. Due to technical limitations of OGM, cases with pathogenic single-nucleotide variants (SNVs) and small insertions and deletions (Indels) identified by sequencing, Robertsonian translocations, balanced centromeric translocations, and mosaicism below 20% cellular fraction were excluded from this study.^8^

This cohort consists of 219 de-identified independent cases (Figure 1) with various clinical indications including abnormal ultrasound findings, positive noninvasive prenatal testing (NIPT), advanced maternal age, family history, etc. Cases were assessed by one or more conventional cytogenetic tests, including CMA, FISH and karyotype (Figure 2A) and enriched for a variety of known chromosomal aberrations (Figure 1 and 2B). CMA platforms included Agilent Technologies (Santa Clara, CA, USA) 180K Oligonucleotide + SNP Array CGH, Illumina (San Diego, CA, USA) CytoSNP-850Kv1.2 BeadChip, and Affymetrix (Santa Clara, CA, USA) Cytoscan HD.

**Figure 1.**
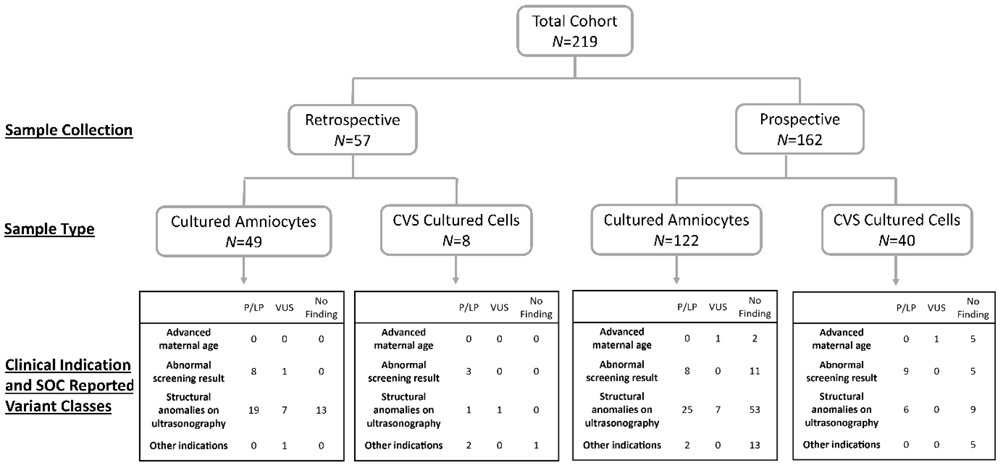
Cohort characteristics, clinical indications, and SOC reported variants. P/LP - pathogenic or likely pathogenic; VUS – variant of uncertain significance.

**Figure 2.**
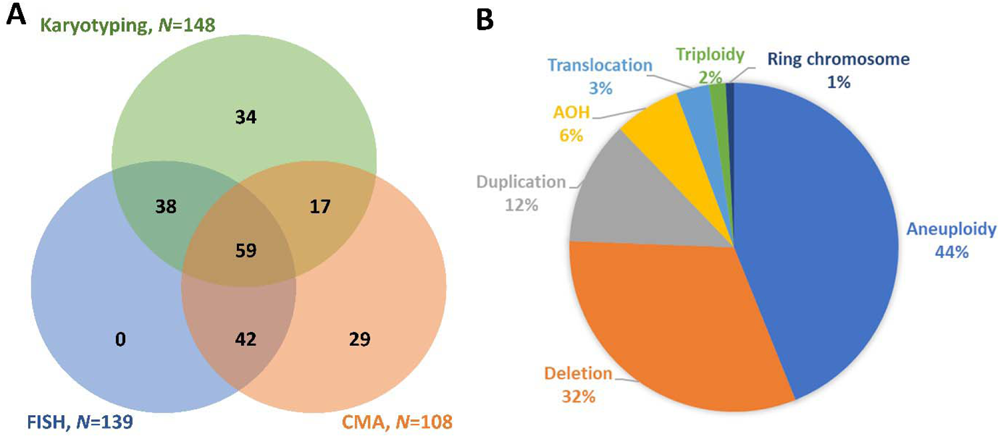
SOC tests and reported variant type. **A.** Distribution of cases having one or more SOC test performed. **B.** Variant types reported by SOC test.

### Sample preparation and processing

Preparation and processing of samples were similar to the initial phase of the study.^11^ Sample contributing sites prepared aliquots of cultured and cryopreserved amniocytes or cultured cells from chorionic villus samples (CVS). Cells cultured in flasks were detached with trypsin-EDTA, rinsed into conical tubes and pelleted, counted, and cryopreserved in freezing medium with 5% DMSO targeting 1 – 1.5 million viable cells per sample. Anonymized samples were sent to a central accessioning site and then distributed to sites enrolled as OGM data collection labs.

Ultra-high molecular weight (UHMW) DNA was isolated using Bionano Prep® SP or SP-G2 kits and labeled using Bionano Prep® DLS or DLS-G2 kits (Bionano, San Diego, CA, USA). Briefly, cryopreserved samples were first thawed and counted. Subsequently, samples were subjected to a series of steps including washing, pelleting from the supernatant, resuspension, and digestion using Proteinase K, RNase A, and lysis buffer. The resulting DNA was then precipitated with isopropanol and bound to a nanobind magnetic disk. After several washing steps, DNA was eluted into a buffer overnight. Solubilized DNA was measured for its concentration. 750 ng DNA was then labeled at a 6-base pair motif (CTTAAG) utilizing a Direct Label and Stain assay. The labeled DNA solution underwent quantification for quality control. Finally, it was loaded onto Saphyr chips for imaging. In the imaging stage, fluorescently labeled DNA molecules were sequentially visualized as they traversed nanochannel on a Saphyr instrument. The target for data collection aimed to reach 800 gigabases (Gbp) for each sample.

### Assay quality control

The evaluation of UHMW DNA quality includes observation of viscosity followed by assessing the DNA concentration and homogeneity (target range 36-150 ng/μl and CV for triplicate measurements < 30%). The first pass success rate, which includes DNA isolation, labeling, and Saphyr chip run, was determined by the data output meeting predefined Quality Control (QC) criteria during the initial assay run. The final assay success rate was assessed based on the data output meeting these QC criteria over one or more rounds of the assay.

A Molecule Quality Report (MQR) was generated for each dataset, which included three essential metrics for sample QC evaluation: Molecule N50 (with a criterion of ≥150 kbp), map rate, and effective coverage. Molecule N50 was used to assess the size distribution of DNA molecules with lengths greater than or equal to 150 kbp. The map rate was calculated as the proportion of DNA molecules that successfully aligned to the GRCh38 reference genome. Effective coverage was calculated based on the depth of coverage of molecules aligned to the same reference genome. Analytical QC targets were defined to achieve: an N50 of ≥230 kbp for molecules ≥150 kbp, a map rate of ≥70%, and an effective coverage of the genome (GRCh38) of ≥160 times.

Post-analytical quality control performance was evaluated by determining the sex of each sample and measuring performance at stable regions of the genome. The EnFocus™ Fragile X Analysis pipeline was used to infer the sample’s sex and establish pass/fail criteria for post-analytical quality metrics (Figure 3). Stable regions, one from each autosome, were identified as regions with minimal variation in control populations. The absolute percent differences between the OGM map and the reference genome should not exceed 1.2% based on expected sizing errors. The pipeline required that at least 90% of the stable regions meet this threshold to pass the quality control criteria.

**Figure 3.**
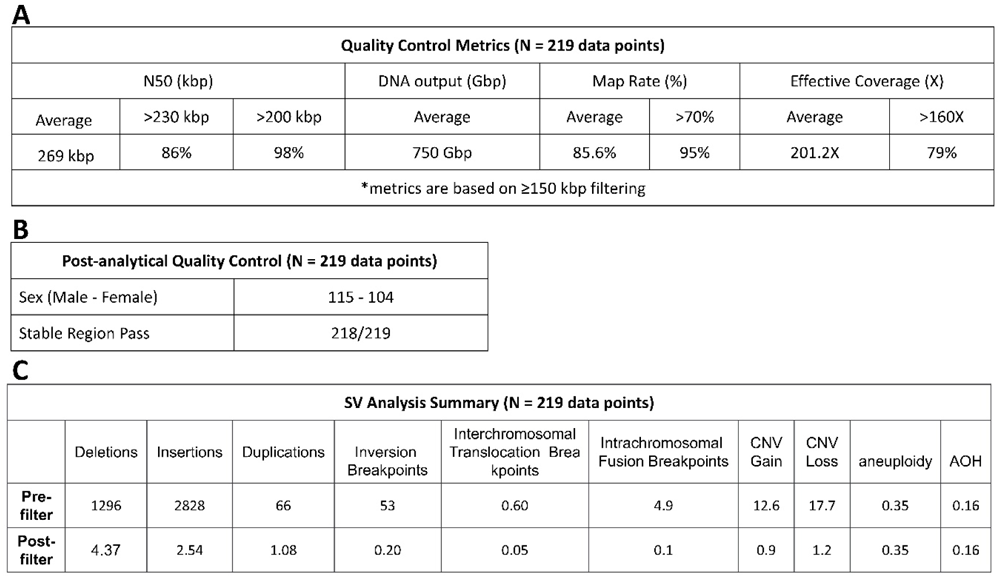
OGM quality control metrics **A.** OGM quality control metrics based on molecules ≥150 kbp. **B.** Post-analytical quality control metrics of gender and stable region check. **C.** SV analysis summary for the baseline average before filtering and after standardized filtering.

### Data analysis

Genome-wide SV analysis in this study was conducted using Bionano Solve™ software version 3.7 and Bionano Access™ software version 1.7. Structural variants (SVs) were identified based on the alignment between the de novo assembled genome maps and the GRCh38 reference. Fractional copy number (CN) analyses were conducted by aligning molecules and labels against GRCh38. Raw label coverage from a sample was normalized, segmented, and baseline CN states were estimated by calculating the mode of coverage for all labels. In cases where chromosome Y molecules were present, baseline coverage in sex chromosomes was halved. With the baseline established, CN states of segmented genomic intervals were evaluated for significant increases or decreases compared to the baseline. Aneuploidies were determined by comparing normalized coverage between chromosomes, and the absence of heterozygosity (AOH) was identified when segments with SV homozygosity exceeded expected values, specifically when larger than 25 Mbp.

### SV interpretation

The analysis of variants was conducted using Bionano Access™ software version 1.7. These variants went through a two-step filtering process. First, they were subjected to a default set of baseline quality control filters, which included the recommended confidence and size thresholds from Bionano Access v1.7. Subsequently, laboratory analysts applied a defined set of filters to prioritize the most relevant variants. The criteria for the filters were as follows: variants with a population frequency exceeding 1% in an OGM-specific controls database (over 300 phenotypically healthy individuals), variants that did not overlap with genes in the GRCh38 reference within a 3-kilobase pair (kbp) range, and deletions and insertions smaller than 1.5 kbp were excluded from consideration. Conversely, SVs and CNVs that overlapped with masked regions of GRCh38 were included in the analysis. Following these filtration steps, laboratory analysts curated a list of variants and then specifically searched for translocation and intra-fusion (large-scale >5Mbp on same chromosome) breakpoints with the GRCh38 gene overlap criterion removed.

Variants that successfully passed the filtration steps underwent analysis and classification based on the guidelines established by the American College of Medical Genetics and Genomics.^12^ Classification categories included pathogenic, likely pathogenic, variant of uncertain significance (VUS), likely benign and benign. This classification process was performed in a blinded manner within Bionano Access by a laboratory analyst and a certified laboratory director. Additionally, large-scale abnormalities such as aneuploidies, triploidy, or regions of absence of heterozygosity (AOH) that were not integrated into the curation and classification process described above were included in the final summary.

### Concordance assessment

Assessment for concordance between OGM data and the SOC results was carried out by a small analyst team that was not part of the blinded clinical interpretation process. The evaluation of variant concordance was conducted through a combination of methods, including software-based calling, visual inspection, and manual curation. For cases without a genetic diagnosis, technical concordance was assessed by checking whether the samples exhibited any significant events that should have been detectable by the respective negative SOC method.

Analytical performance metrics were assessed by using 123 aberrations that were reported in SOC testing (Table 1). Analytical sensitivity, specificity, and positive predictive value (PPV), negative predictive value (NPV), and accuracy were evaluated. Cases without SOC reported findings were used for false-positive and true-negative calculations.

**Table 1.** Analytical Performance Metric Evaluation.

| Performance criteria | Overall Variants (n = 123) | Aneuploidy (n = 54) | Deletion (n = 39) | Duplication/gains (n = 15) | AOH (n = 8) | Translocation (n = 4) | Triploidy (n = 2) | Ring chromosome (n = 1) |
| --- | --- | --- | --- | --- | --- | --- | --- | --- |
| Sensitivity, % | 99.2 | 100 | 100 | 100 | 100 | 100 | 100 | NA* |
| Specificity, % | 100 | 100 | 100 | 100 | 100 | 100 | 100 | NA* |
| PPV/precision, % | 100 | 100 | 100 | 100 | 100 | 100 | 100 | NA* |
| NPV, % | 95.5 | 100 | 100 | 100 | 100 | 100 | 100 | NA* |
| Accuracy, % | 99.6 | 100 | 100 | 100 | 100 | 100 | 100 | NA* |

Sensitivity/positive percentage agreement = true positive/(true positive + false negative).
Specificity/negative percentage agreement = true negative/(true negative + false positive).
PPV = true positive/(true positive + false positive).
NPV = true negative/(true negative + false negative).
Accuracy = true positive + true negative/all results.
123 SOC reported variants from 102 individuals were used for true positive and false negative; 117 cases without SOC reported findings were used for false positive and true negative calculations.
PPV, positive predictive value; NPV, negative predictive value; SV, structural variation; NA, not available.
\* The mosaic ring chromosome X in Case-036 is not called by OGM.

## Results

### Cohort description and SOC testing

Two hundred and nineteen cases underwent OGM workflow and data analysis. 57 cases were retrospective cases and 162 cases were prospective cases. Forty-nine retrospective cases were cultured amniocytes and eight were CVS cultured cells, while 122 prospective cases were cultured amniocytes and 40 were CVS cultured cells (Figure 1). Indications for prenatal diagnosis are categorized as advanced maternal age (*n*=9), abnormal screening result (*n*=45), structural anomalies on ultrasonography (*n*=141), and other indications (*n*=24). Overall, there were 82 cases with reported pathogenic findings, one case with reported likely pathogenic finding, 19 cases with reported VUS, and 117 cases without a reported finding. More details for SOC reported variant classification and breakdown of clinical indications are shown in Figure 1.

Because there are several prenatal diagnostic options available and each method has its strengths and limitations, multiple tests were often ordered concurrently or consecutively in the clinical settings. In this cohort, more than one conventional SOC test was performed in 71.2% of the cases (156/219). There were 19.2% (42/219) of cases with FISH and CMA, 17.4% (38/219) of cases with karyotyping and FISH, 7.8% (17/219) with karyotyping and CMA, and 26.9% (59/219) with all three of these SOC methods (Figure 2). Breakdown of the variant type reported by SOC tests is shown in Figure 2B and more details are provided in the Supplementary Table 1. There were 123 SOC reported findings, and the majority were aneuploidy (44%), followed by deletion (32%), duplication (12%), AOH (6%), translocation (3%), triploidy (2%), and ring chromosome (1%).

### Data quality control and SV summary

Overall data quality of this cohort exceeded the baseline. Average DNA output was 750 Gbp, with the average molecule size (N50≥150 kbp) at 269 kbp. The average map rate was at 85.6%, and 95% of the cases had a map rate greater than 70%. The average effective coverage was at 201.2X and there were 79% of cases having coverage greater than 160X (Figure 3A).

Post-analytical quality control revealed that there were 115 male and 104 female samples, which was consistent with information gathered during sample intake. For the stable region check, 218 out of 219 samples passed the quality check (Figure 3B). Only one case, Case-076, failed stable region QC check. After reviewing the overall data quality of Case-076, this sample was determined to be analyzable and subsequently proceeded to further analysis.

On average, 4,279 SVs were initially identified in each case in the whole genome analysis. After applying filtering criteria described in the Methods, an average of 11 SVs were found in each case which underwent classification by analysts and directors based on ACMG guidelines (Figure 3C).

### Concordance and Analytical Performance Metric Evaluation

OGM data were compared to SOC test results to determine concordance, and this resulted in 100% concordance from 219 clinical cases in this cohort (Supplementary Table). One female case (Case-036) was determined to be partially concordant. Based on karyotyping (45,X[8]/46,X,r(X)(p11.23q21.3)[7]) and CMA (arr[GRCh38] Xp22.33p11.3(10,814-46,826,640)x1, Xp11.3q21.31(46,837,624-89,823,554)x1∼2, Xq21.31q28(89,922,674-156,007,082)x1) findings, this case had monosomy of chromosome X and a mosaic ring chromosome X. Monosomy of chromosome X was successfully detected by OGM; however, the mosaic ring chromosome X was not detected in this case. To investigate the possibility of losing mosaic ring chromosome X during cell culturing of amniocytes, extracted UHMW DNA was sent for orthogonal CMA test for confirmation, which is still pending.

Analytical performance metric was evaluated using 123 SOC reported variants (82 pathogenic, one likely pathogenic and 19 VUS) from 102 individuals (Supplementary Table 1). Overall, 122 out of 123 variants were detected by OGM and the mosaic ring chromosome X is the only variant missed by OGM. Although the presence of the mosaic ring chromosome X in UHMW DNA of Case-036 is not yet confirmed, it was counted as false negative in this analysis.

Therefore, OGM have sensitivity of 99.2%, specificity of 100%, positive predictive value (PPV) of 100%, negative predictive value (NPV) of 95.5% and accuracy of 99.6% (Table 1). Performance broken down by cytogenomic variant type is detailed in Table 1.

## Discussion

Global medical associations, including the American College of Obstetrics and Gynecology (ACOG), the International Society of Ultrasound in Obstetrics and Gynecology (ISUOG), and the American College of Medical Genetics and Genomics, recommend prenatal genetic screening for all pregnant individuals, regardless of gestational or maternal age.^13–15^ In instances where NIPT yields a positive result or when there’s an inconclusive outcome due to technological constraints, further diagnostic testing is recommended to validate the screening findings. Moreover, pregnancies exhibiting abnormal ultrasound findings indicating fetal defects are advised to undergo further diagnostic invasive procedures, such as CVS, amniocentesis, or periumbilical blood sampling.^16^ Historically, confirmatory diagnostic procedures utilized conventional cytogenetic methods like karyotyping and FISH.^17–18^ However, advancing into the 2000s, the introduction of CMA using amniocytes or CVS emerged as the preferred initial diagnostic tool recommended by ACOG and ISUOG for pregnancies considered high-risk for fetal genetic disease and those with abnormal ultrasound results.^19–20^ This shift marked a more comprehensive approach to genetic profiling, providing enhanced insights into suspected fetal genetic anomalies.

OGM has the capability to identify a wide range of chromosomal anomalies, spanning aneuploidies, sex chromosome variations, microdeletions/microduplications, CNVs, balanced chromosomal occurrences, and complex chromosomal rearrangements. Compared to conventional cytogenetic techniques, OGM offers a higher resolution in detecting these aberrations. As a next-generation cytogenomic tool, OGM holds promising potential in prenatal care and management due to its enhanced capacity for comprehensive chromosomal analysis.^7^

This ongoing multisite study focuses on assessing and validating OGM analysis in comparison to established SOC methodologies in prenatal assessment. Phase 1 findings from this study revealed robust results, demonstrating high concordance, accurate variant detection, and reproducibility across multiple participating sites.^11^ This report outlines the advancements and developments observed in the ongoing phase 2 of the study. The focus remains on evaluating the efficacy and comparative merits of OGM against established SOC practices in prenatal diagnosis.

The phase 2 cohort comprises 219 retrospective and prospective cases (Figure 1). Within this cohort, a substantial portion has undergone multiple conventional SOC tests. Notably, 71% of these cases have been subjected to more than one conventional SOC examination, while 27% have undergone the complete trio of SOC tests, including karyotyping, FISH, and CMA. This highlights the strength and limitation of each SOC test; consequently, these tests are frequently employed either in tandem or simultaneously in prenatal clinical settings.

The data collected throughout phase 2 of the study exceeded the initial QC baseline (Figure 3A). All cases yielded analyzable data, including a case (Case-076) that initially did not meet the stable region QC criteria (Figure 3B). The subsequent variant calling resulted in 100% concordance between OGM and the SOC methodologies. Among the 219 cases assessed, 218 demonstrated complete concordance between OGM and SOC, while a female case (Case-036) exhibited partial concordance in the study. In Case-036, karyotyping and CMA revealed monosomy of chromosome X and a mosaic ring chromosome X (estimated 40-50% cellular fraction). While OGM successfully detected the monosomy of chromosome X, it did not identify the presence of the mosaic ring chromosome X in this case. Considering the potential for the loss of mosaic ring chromosome X during the cell culturing process of amniocytes, extracted UHMW DNA was sent for orthogonal CMA testing to confirm the presence of the ring chromosome X. However, the result is still pending during the writing of this report. This highlights the robustness and reliability of data collection, as well as high concordance in comparison to SOC.

Evaluation of analytical performance metrics was conducted by utilizing 123 variants reported by SOC. OGM demonstrated high performance metrics: 99.2% sensitivity, 100% specificity, 100% PPV, 95.5% NPV, and an overall accuracy of 99.6% (Table 1). These results are consistent with the performance observed in the earlier phase 1 study.^11^ The consistent high performance metrics demonstrate OGM’s reliability and its ability to effectively detect abnormalities identified by SOC testing of prenatal samples.

## Conclusions

The study demonstrated the potential of OGM as a high-resolution cytogenomic assay suitable for application after an abnormal NIPT screen or in high-risk pregnancies with abnormal ultrasound results. Multiple studies, including this one highlight OGM’s performance and its distinctive capability to identify all categories of clinically relevant genome-wide structural variations (SVs) at high resolution. This update on phase 2 progress contributes to the expanding evidence base, emphasizing OGM’s promise as a stand-alone and promising alternative assay in prenatal diagnosis, distinct from traditional cytogenetic methods.

To date, a total of 342 unique cases (419 data points) in this ongoing multisite prenatal clinical study have been reported through this publication and Stevenson et al., 2022.^11^ The goal is to comprehensively document and report findings from the entire cohort, expected to exceed 500 unique cases during the year 2024.

## Data Availability

Data will be made available upon reasonable request and in accordance with IRB protocols.

## Acknowledgements

The authors wish to acknowledge Bionano clinical affairs team: Alka Chaubey, Alex Hastie, Mike Gallagher, James Yu, Ben Clifford, Natacha Diaz-Meyer, Stephanie Burke, Andy Pang, Jen Hauenstein, Alex Chitsazan, Joey Estabrook, Kelsea Chang, Shuk Shukor, and Ileana Carrera for support in training and data management; and Bionano Laboratories: Trilochan Sahoo for support in discussions.

